# Multimodal Deep Learning Model to Estimate CT-based Body Composition Measures Using Chest radiographs and Clinical Data

**DOI:** 10.1101/2025.01.16.25320684

**Authors:** Ehsan Alipour, Samuel Gratzl, Ahmad Algohary, Hao Lin, Manoj Bapat, Duy Do, Charlotte Baker, Tricia Rodriguez, Brianna Cartwright, Jennifer Hadlock, Peter Tarczy-Hornoch, Anand Oka, Nicholas Stucky

**Affiliations:** Truveta Inc, Bellevue, WA; Department of Biomedical and Health Informatics, University of Washington, Seattle, WA; Institute for Systems Biology, Seattle, WA

## Abstract

Body composition metrics such as visceral fat volume, subcutaneous fat volume and skeletal muscle volume, are important predictors for cardiovascular disease, diabetes, and cancer prognosis. Recent advances in artificial intelligence have enabled automatic calculation of body composition metrics from CT scans and MRIs.

In this study, we explore using deep learning to estimate body composition metrics from chest radiographs and a small set of variables that are easy to obtain in the clinical environment.

A retrospective cohort of patients with concurrent non-contrast abdominal CT scan and chest radiograph was selected using the Truveta dataset. TotalSegmentator was used to delineate various tissue components in the CT scan. Volumetric and mid-l3 level body composition measures including subcutaneous and visceral fat volume, skeletal muscle and bone related indices, and aortic calcification scores were calculated. A multitask, multimodal deep learning model using chest radiographs and select clinical variables was trained to estimate the ground truth body composition metrics. Three data fusion strategies were compared in this process: early, intermediate, and late.

Our final cohort consisted of 1,118 patients. Mean age of imaging was 67 years old, mean height was 1.67 meters and mean weight was 78 kgs. The late fusion multimodal model outperformed both unimodal clinical-only and imaging-only models, in addition to the early and intermediate fusion models. It achieved a Pearson correlation of 0.85 in prediction of subcutaneous fat volume and 0.76 and 0.72 in estimating visceral fat volume and vertebral bone volume in the hold-out test set.

While previous studies have used artificial intelligence to automate calculation body composition metrics in CT scans and MRIs, we introduce a novel strategy that utilizes simpler imaging modalities. Our models can be used in future studies to calculate body composition metrics for larger cohorts of patients using only a chest radiograph and readily available clinical variables.

## Introduction

Body composition metrics have attracted significant attention as predictors of health-related outcomes. Multiple studies have demonstrated correlations between body composition metrics like visceral fat area and skeletal muscle index and chronic non-communicable disease including cardiovascular disease and diabetes (1,2). In addition, studies have found that abnormal body composition measures can be a predictor of poor prognosis in patients with cancer including those with breast and colorectal cancers(3). These correlations are often stronger than correlation between these diseases and commonly used body composition surrogates like body mass index (BMI) and weight (1,2,4). Measures such as BMI fail to capture critical information about body composition including muscle mass, visceral adipose tissue (which has been shown to be the associated with pathologic conditions (5)) and subcutaneous adipose tissue. Studies have also explored the use of body composition metrics acquired through CT scan or MRI for opportunistic screening of various diseases including cardiometabolic disease, osteoporosis, and steatohepatitis (6).

Common methods for calculating body composition metrics include whole body magnetic resonance, computed tomography imaging or dual-energy x-ray absorptiometry (DXA), and bioelectrical impedance analysis (7). However, these methods may not be available for all patients due to limitation of resources, risk of radiation exposure, or need for specialized equipment, software and staff. Scientists have validated using body composition metrics calculated from 2D slices in the L3 section of abdominal CT scans as surrogates of whole-body composition metrics (8,9). While CT and MRI provide accurate estimates of body composition (4), performing a CT scans or MRI to calculate body composition metrics on the general public for opportunistic screening poses challenges, including high costs and risk from higher radiation exposure(4,7). Hence, new strategies to estimate body composition measures using readily accessible clinical and imaging data would enable calculation of body composition metrics for a larger pool of individuals. One promising avenue is to use simpler modalities like chest radiographs to estimate body composition. A recent study on opportunistic screening of type 2 diabetes demonstrated that chest radiographs encode information about fat distribution that can be used to screen for type 2 diabetes (10).

In this study, we aim to create a multi-modal deep learning model to estimate CT-based body composition metrics by combining readily available clinical data with chest radiographs. Such a model can be applied with minimal cost retrospectively or prospectively on any person with a chest radiograph, enabling greater use of body composition metrics in research, and clinical care including screening. In addition, we will experiment with different ways of combining the clinical data and imaging data.

## Methods

To ensure that our study meets the requirements of high-quality clinical AI research, we followed the checklist of artificial intelligence in medical imaging (CLAIM) guidelines in all steps of our study (11).

### Dataset

The clinical and imaging data used in our study was compiled from the Truveta Data. Truveta provides access to continuously updated and linked EHR data. It consisted of deidentified records from a subset of the Truveta Data which also contributed imaging data. Truveta Data used in this study was accessed on July 14^th^, 2024. This study used only de-identified patient records and therefore did not require Institutional Review Board approval. Refer to **supplementary material section 1** for additional details.

Using Truveta Data, we identified adult individuals who had a chest radiography within 3 months of an abdominal CT scan without any contrast agent. We further limited our inclusion criteria to patients with a weight measurement within 1 month of the chest radiography and had a height measurement anytime in their adult life. Although our search returned 312,444 unique cases, due to computational constraints, we randomly sample 3,000 patients. The imaging data, in addition to the age at the time of imaging, sex at birth, weight, and height were collected for every patient. Additional details are available in **supplementary material section 1**. The cohort flowchart diagram can be seen in **Figure 1**.

**Figure 1.**
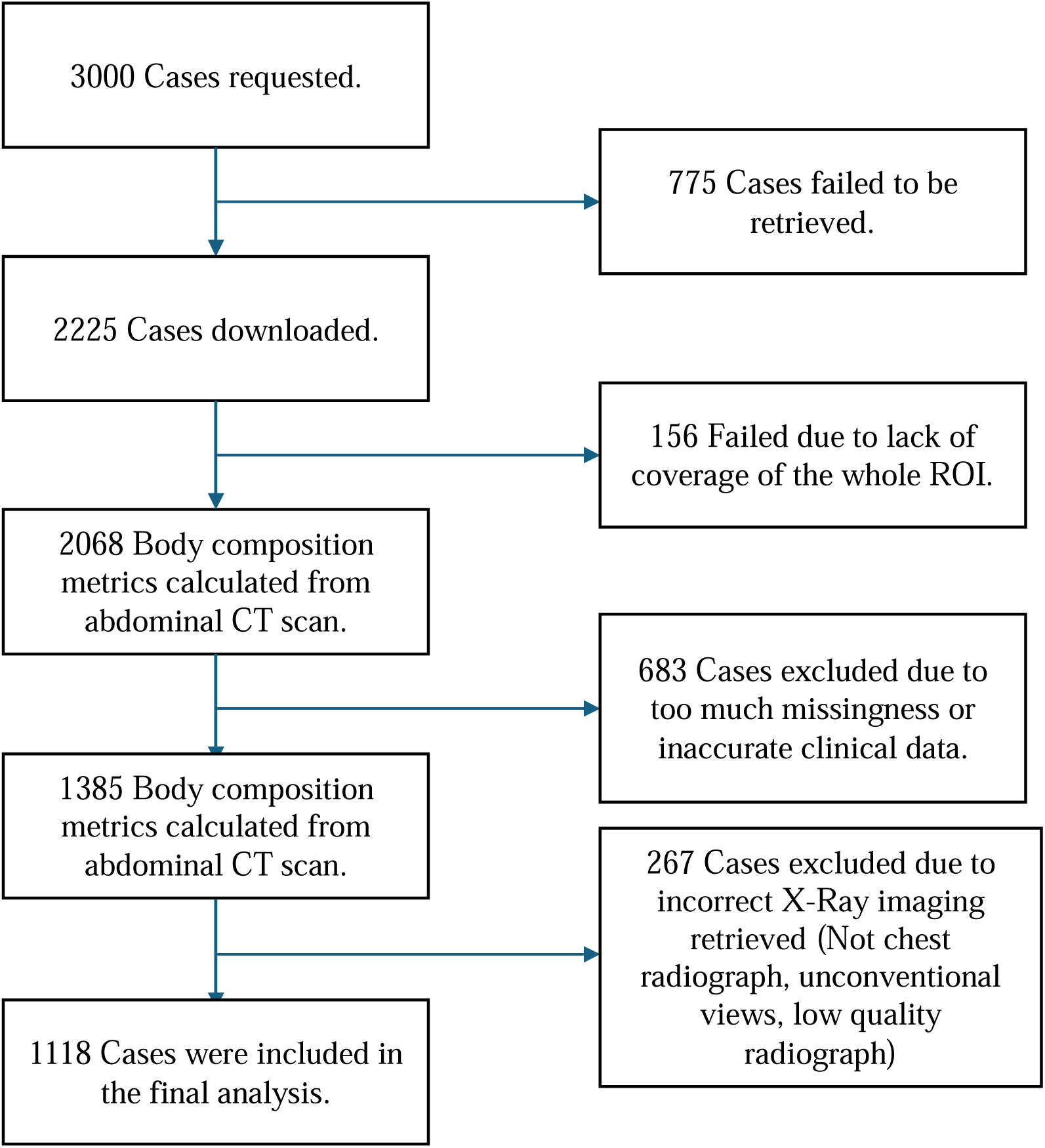
Cohort Flowchart. We initially request 3000 cases. Out of these, 1118 cases met all our inclusion criteria and were included in our study. ROI: Region of Interest.

### Missing Data

For cases that the recorded height or weight were outside of acceptable range or were missing, we imputed the missing value using linear regression on the rest of the variables.

### Body Composition Metrics Calculation

The ground truth values for body composition were calculated using a combination of 2D slices from the T12 vertebrae to the L5 vertebrae for volumetric body composition metrics and on the mid-L3 level for the single slice body composition metrics in the abdominal CT scans. The TotalSegmentator (12) tool was used to segment out the various body composition sections in CT scans and identify the T12, L3 and L5 levels. The list of body composition metrics that were calculated can be found in **Table 1** (for L3-level body composition metrics, refer to **Table S1** in the appendix.). An in house rule based python script based on available literature was used to calculate the body composition metrics from the segmentation masks (13–15). More details can be found in **supplementary material section 1**.

**Table 1.** Population Characteristics and Volume Based Body Composition Distribution Across the Training, Validation and Test sets. L3-based body composition distribution is available in the appendix.

|  |  | Train<br>(n = 726) |  | Validation<br>(n = 177) |  | Test<br>(n = 215) |  | All<br>(n = 1118) |  | Significance |
| --- | --- | --- | --- | --- | --- | --- | --- | --- | --- | --- |
|  |  | mean | std | mean | std | mean | std | mean | std | P-value |
| Variable | Category |  |  |  |  |  |  |  |  |  |
| Age |  | 67.13 | 17.36 | 68.15 | 16.31 | 66.13 | 16.8 | 67.1 | 17.09 | 0.51 |
| Height<br>(Meters) |  | 1.69 | 0.18 | 1.66 | 0.16 | 1.7 | 0.27 | 1.69 | 0.2 | 0.12 |
| Weight (Kilograms) |  | 77.34 | 19.66 | 79.66 | 22.11 | 77.74 | 20.28 | 77.78 | 20.18 | 0.39 |
|  |  | Count | % | Count | % | Count | % | Count | % |  |
| Sex | Female | 374 | 51.52 | 90 | 50.85 | 118 | 54.88 | 582 | 52.06 | 0.64 |
|  | Male | 352 | 48.48 | 87 | 49.15 | 97 | 45.12 | 536 | 47.94 |  |
|  |  | mean | std | mean | std | mean | std | mean | std | P-value |
| Skeletal Muscle Volume (cm<sup>3</sup>) |  | 2241.42 | 783.94 | 2265.02 | 848.56 | 2233.83 | 843.84 | 2243.69 | 805.52 | 0.92 |
| Visceral Fat Volume (cm<sup>3</sup>) |  | 2850.55 | 1940.01 | 2877.18 | 1734.09 | 2791.08 | 1916.73 | 2843.33 | 1902.95 | 0.89 |
| Subcutaneous Fat Volume (cm<sup>3</sup>) |  | 4464.93 | 2832.39 | 4789.71 | 2782.95 | 4560.83 | 2700.3 | 4534.79 | 2799.63 | 0.38 |
| Vertebral Bone Volume (cm<sup>3</sup>) |  | 347.09 | 76.06 | 347.6 | 85.02 | 342.74 | 73.76 | 346.34 | 77.06 | 0.75 |
| Fat Free Volume (cm<sup>3</sup>) |  | 7982.08 | 2166.6 | 8249.24 | 2807.68 | 8137.64 | 2421.02 | 8054.29 | 2328.64 | 0.33 |
| Intramuscular Fat (cm<sup>3</sup>) |  | 220.17 | 167.36 | 213.24 | 156.64 | 224.87 | 175.39 | 219.98 | 167.18 | 0.79 |
| Vertebral Bone Density (HU) |  | 320.87 | 78.46 | 322.87 | 71.53 | 329.11 | 86.03 | 322.77 | 78.93 | 0.4 |
| Muscle Radiodensity (HU) |  | 19.49 | 15.4 | 19.52 | 13.29 | 19.23 | 15.14 | 19.44 | 15.02 | 0.97 |
| Aortic Calcification Score |  | 8.67 | 14.58 | 9.22 | 13.97 | 8.72 | 12.55 | 8.77 | 14.11 | 0.9 |
| Number of Plaques in Abdominal Aorta |  | 10.62 | 11.16 | 10.17 | 10.67 | 10.64 | 11.06 | 10.55 | 11.05 | 0.88 |
| Skeletal |  | 792.4 | 276.83 | 828.56 | 326.8 | 788.1 | 303.19 | 797.3 | 290.48 | 0.29 |

| Muscle Index | 3 |  |  | 9 |  |  | 2 |  |  |
| --- | --- | --- | --- | --- | --- | --- | --- | --- | --- |
| Subcutaneous Fat Index | 1620.39 | 1099.3 | 1803.23 | 1198.27 | 1636.58 | 1013.87 | 1652.45 | 1100.84 | 0.14 |
| Visceral Fat Index | 999.76 | 667.44 | 1038.11 | 588.53 | 984.25 | 664.47 | 1002.85 | 654.65 | 0.7 |
| Fat Free Index | 2830.93 | 765.86 | 3026.78 | 1080.12 | 2888.8 | 906.78 | 2873.07 | 852.64 | 0.02 |

**HU:** Hounsfield Units

The data was split with a 0.8/0.2 ratio to generate the training dataset and the hold-out test set. The test set was only used to measure the final model performance. Additionally, 20% of the training data was used as a validation set for hyperparameter tuning. Refer to the **supplementary material section 3** for preprocessing and normalization steps.

### Modeling Approach and Fusion Strategy

The clinical baseline model was developed using a shallow neural network with two fully connected layers. We also developed a regression model using the four clinical variables (height, weight, age, and sex at birth).

The imaging-only model consisted of a multitask convolutional neural network (CNN). Multiple CNN architectures (ResNet10, ResNet50, ResNet101) were tested, however ResNet18 (16) was selected based on validation performance. The network was initiated randomly. The default resnet18 architecture from the PyTorch (v2.4) package was used.

We experimented with three fusion strategies, comparing early, intermediate and late fusion of the clinical and imaging data (17,18). Fusion strategies are depicted in **Figure 2**. More information can be found in the **supplementary material section 2**.

**Figure 2.**
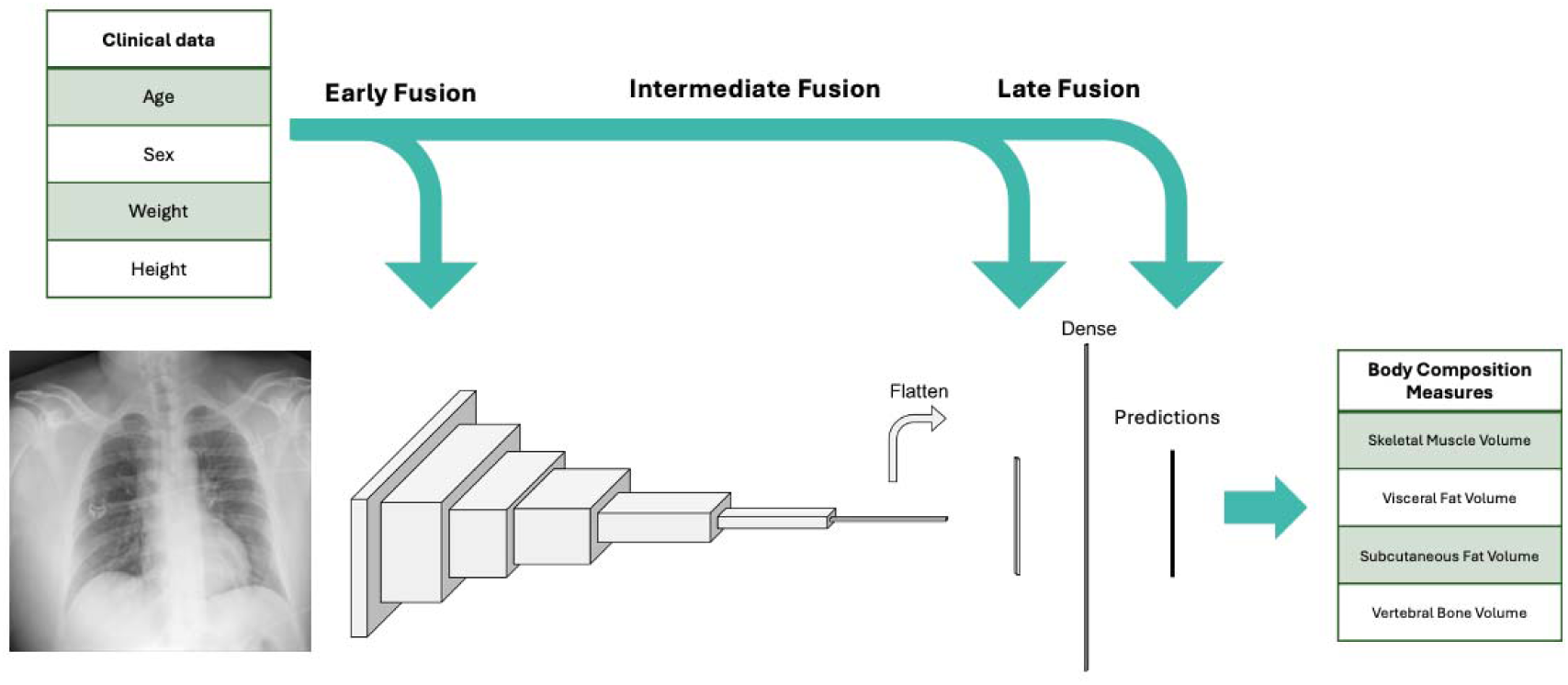
A diagram showing model architecture, input features, and the various fusion strategies used in our study. We used a resnet18 model for our convolutional neural network.

### Evaluation

Huber loss (19), which is a combination of mean absolute error (MAE) and mean squared error (MSE) was used to train the models and evaluate the performance. In addition, weight decay was used for regularization. The correlation between the predicted outcomes versus the ground truth values is reported for all body composition metrics.

### Explainability

Both occlusion sensitivity and integrated gradient methods were used to calculate saliency maps over all the images (20). Aggregate explainability maps were generated by averaging all the input images in the test set and their respective explainability maps for the two methods separately.

### Fairness

Final model performance was measured and compared across the different age, sex and BMI groups present in our test set to calculate performance metrics in different subgroups and identify potential fairness issues with the model.

### Statistical Analysis

P-value significance level threshold was set to 0.05 for all analyses. Pearson correlation was calculated to compare predictions and ground truth values for all body composition metrics except aortic calcification score and number of calcified plaques. Spearman correlation was used for those due to their skewed distribution. ANOVA was used to identify any statistically significant difference present across the data splits for continuous variables and chi square test was used for categorical variables. The following cut points were defined for the correlations: r<0.6 poor, 0.5<r<0.7 moderate, 0.7<r<0.8 good, 0.8<r<0.9 very good and r>0.9 was considered great correlation.

### Data sharing and Code Availability

The data used in this study is available to all Truveta subscribers and may be accessed at studio.truveta.com. Our model weights and the required code to calculate body composition metrics will be released on GitHub at the time of peer-reviewed publication. This project was done using PyTorch (v 2.4), Sci-kit Learn (v. 1.5.2), Captum (0.7.0) on GPU equipped Linux Azure server with one V100 GPU with 16 gigabytes of memory.

## Results

### Study Population

From a potential cohort of 3000 cases, we included a final cohort that consisted of 1118 samples. Some cases were excluded due to problems with identifying the vertebrae levels. Other problems included missingness of both weight and height values, and low-quality chest radiographs, including the image not being a chest radiograph (mislabeled DICOM description), unconventional views, low exposure, or presence of large artifacts in the CXR. The cohort flowchart is shown in **Figure 1**. Average age was 68 ± 17.36 years of age. The average BMI was 27. The demographic distribution of the cases and distribution of the body composition metrics can be seen in **Table 1**.

### Clinical Model

The clinical model included age, sex at birth, height, and weight. The clinical only model achieved good correlation (0.77, 0.71 – 0.82) when predicting subcutaneous fat volume and index but achieved moderate to poor performance in estimating other body composition metrics. The other highest performing metrics included visceral fat area 0.69 (0.65 – 0.71) and vertebral bone volume with a correlation of 0.67 (0.59 – 0.74). More detailed performance metrics can be viewed in **Table 2**.

**Table 2:**
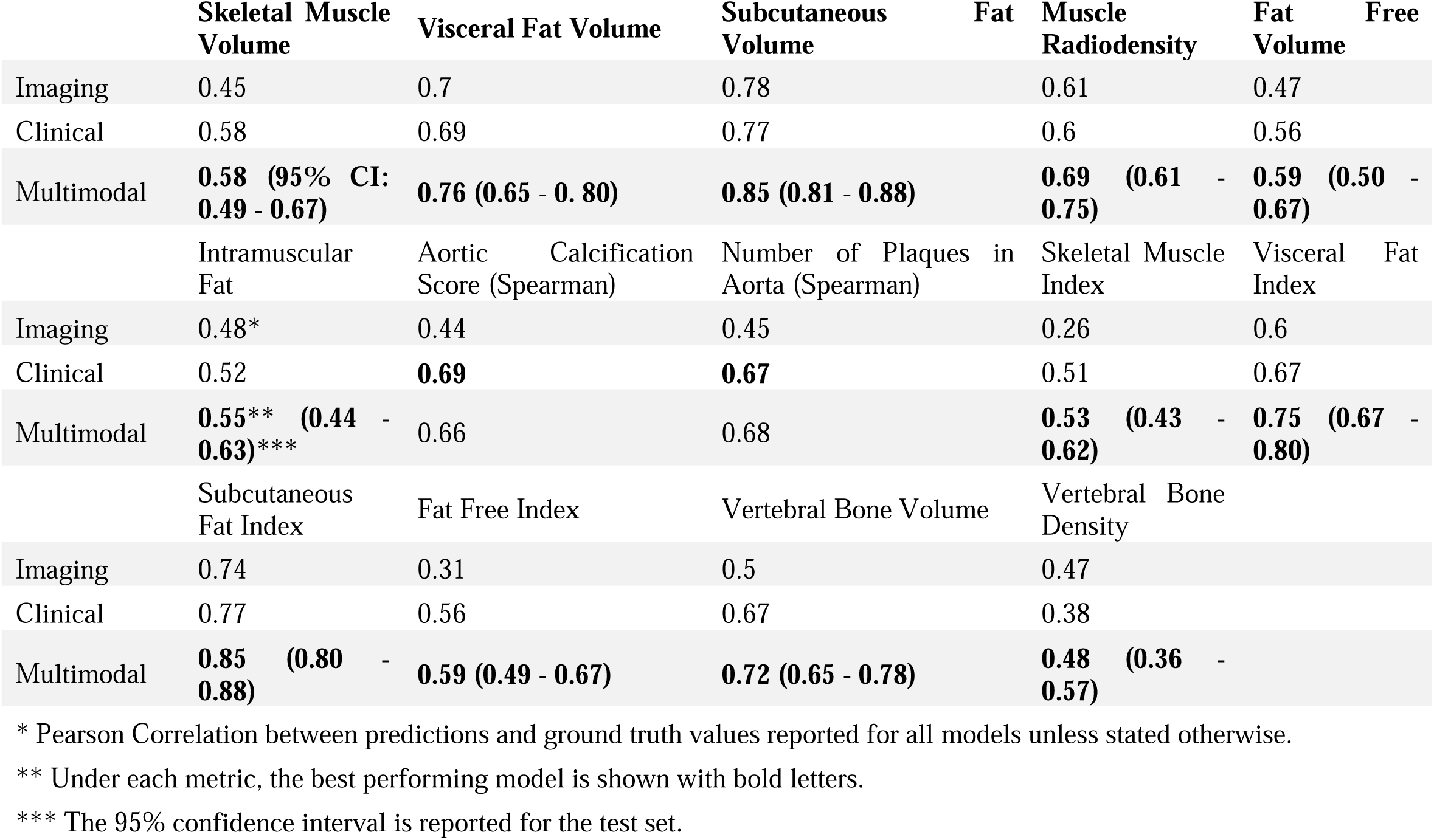
Final model performance in the holdout test set comparing imaging only, clinical only and multimodal models.

### Imaging Model

The image only model outperformed the clinical model in prediction of subcutaneous fat volume (0.78, 0.72 – 0.82) and visceral fat volume (0.70, CI 0.63 - 0.76) achieving good correlation with ground truth in both metrics. The imaging only model also performed better in estimating muscle radiodensity and vertebral bone radiodensity compared to the clinical-only model.

### Combined Model

Our multimodal models achieved higher performance in estimating almost all body composition metrics, especially subcutaneous and visceral fat related measures. Our results showed that late fusion based model performed best in estimating body composition metrics followed closely by intermediate fusion. Scatter plots comparing model performance on the test set for the three top performing body composition metrics could be viewed in **Figure 3**. **Table 3** contains the final multimodal model performance metrics across the train, validation, and test cohorts.

**Figure 3.**
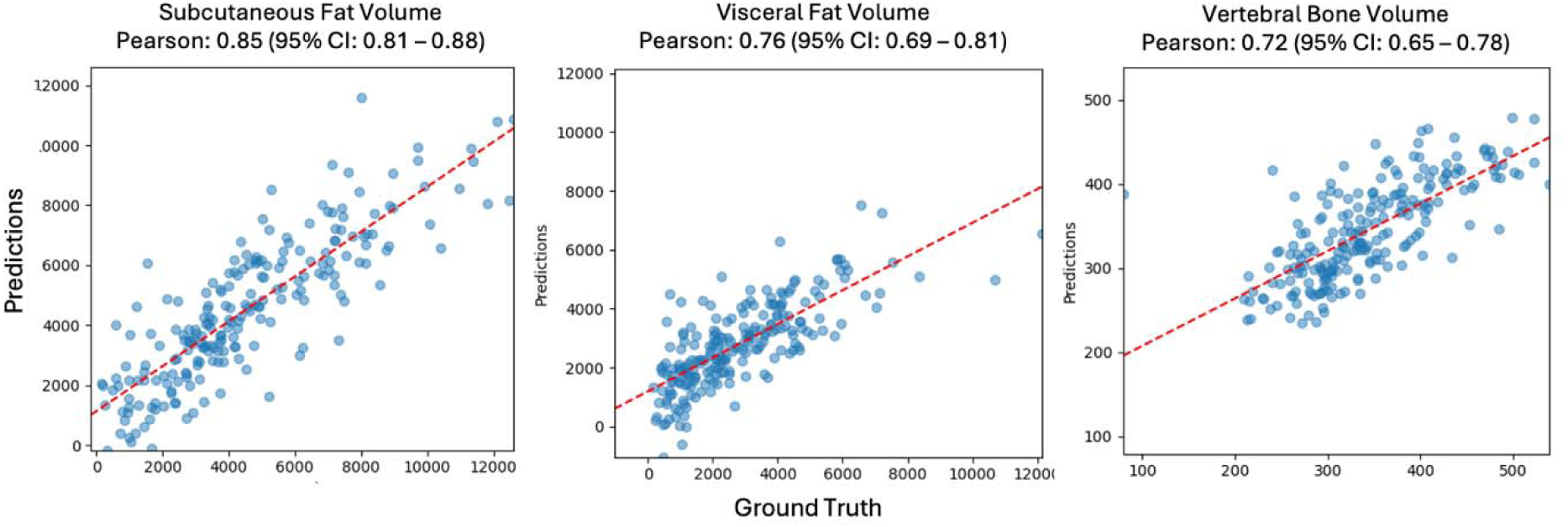
Scatter plots showing multimodal model predictions versus ground truth values for three of the top performing body composition metrics.

**Table 3:**
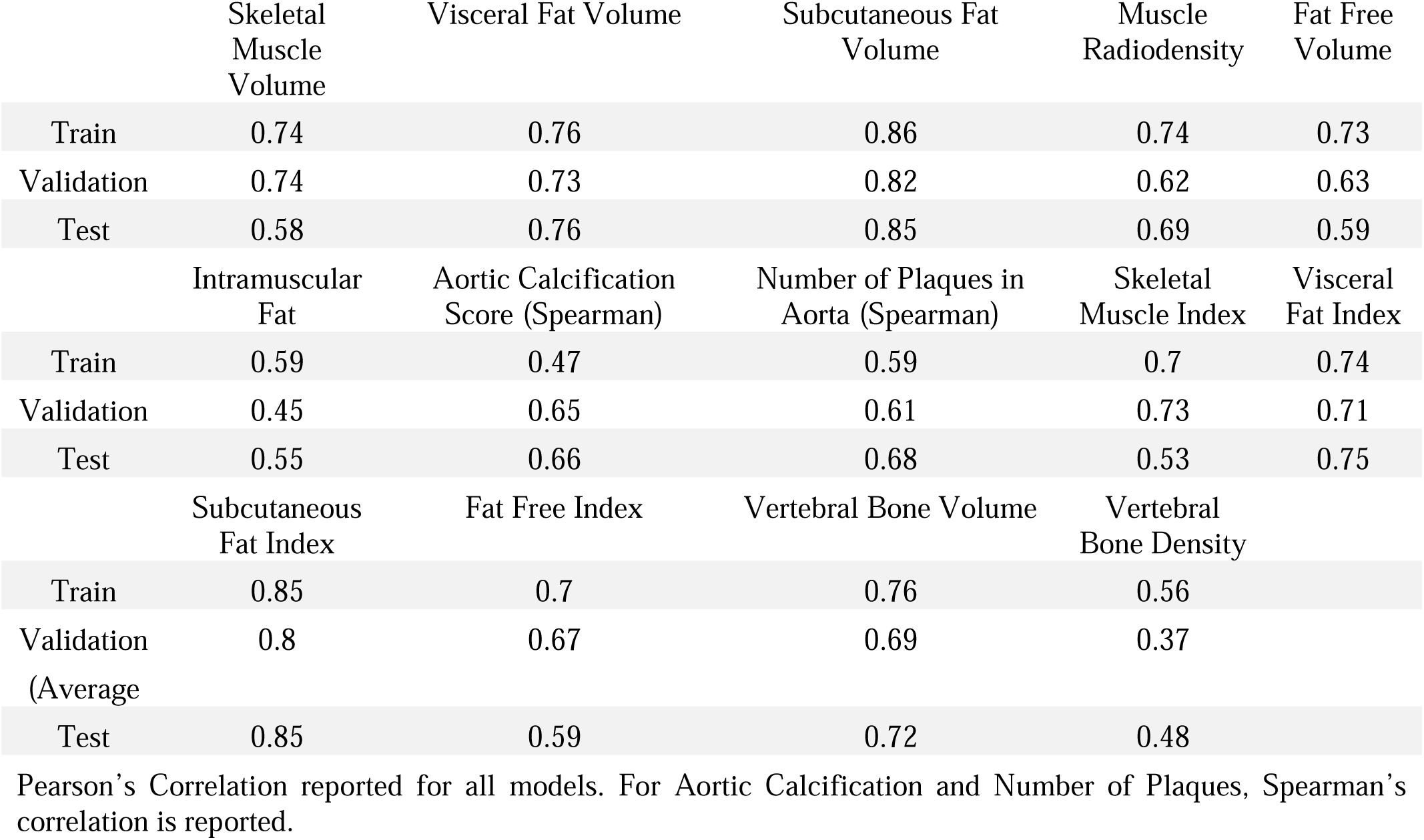
Final multimodal model performance across train, test and validation sets.

#### Early Fusion

The early fusion model estimates achieved good correlation for subcutaneous fat index (0.80, 0.75 - 84) and achieved good correlation for visceral fat index, visceral fat volume and subcutaneous fat volume. The early fusion model consistently underperformed the two other models in estimating all body composition metrics.

#### Intermediate Fusion

The intermediate fusion model achieved a correlation of 0.82 (0.78 – 0.86) in estimating subcutaneous fat volume and index. It also achieved good performance in estimating visceral fat volume and visceral fat index. This model outperformed the other fusion strategies in estimating muscle radiodensity (0.69, 0.61 – 0.75), fat free index (0.60, 0.51 – 0.68), intramuscular fat (0.57, 0.48 – 0.66), and vertebral bone radiodensity (0.49, 0.38 – 0.58).

#### Late Fusion

The late fusion model achieved a correlation of 0.85 (0.81 - 88) in estimating subcutaneous fat volume. This model also had good performance in estimating visceral fat volume (0.76, 0.69 – 0.81) and vertebral bone volume (0.72, 0.65 – 0.78). The Spearman’s correlation for skeletal muscle volume (r = 0.67), vertebral bone volume (r = 0.75) and subcutaneous fat index (r = 0.88) was higher than the Pearson’s correlations highlighting a slightly better ability to rank observations compared to estimating their actual value. **Table 4** contains performance metrics for all body composition measures across the fusion strategies.

**Table 4:**
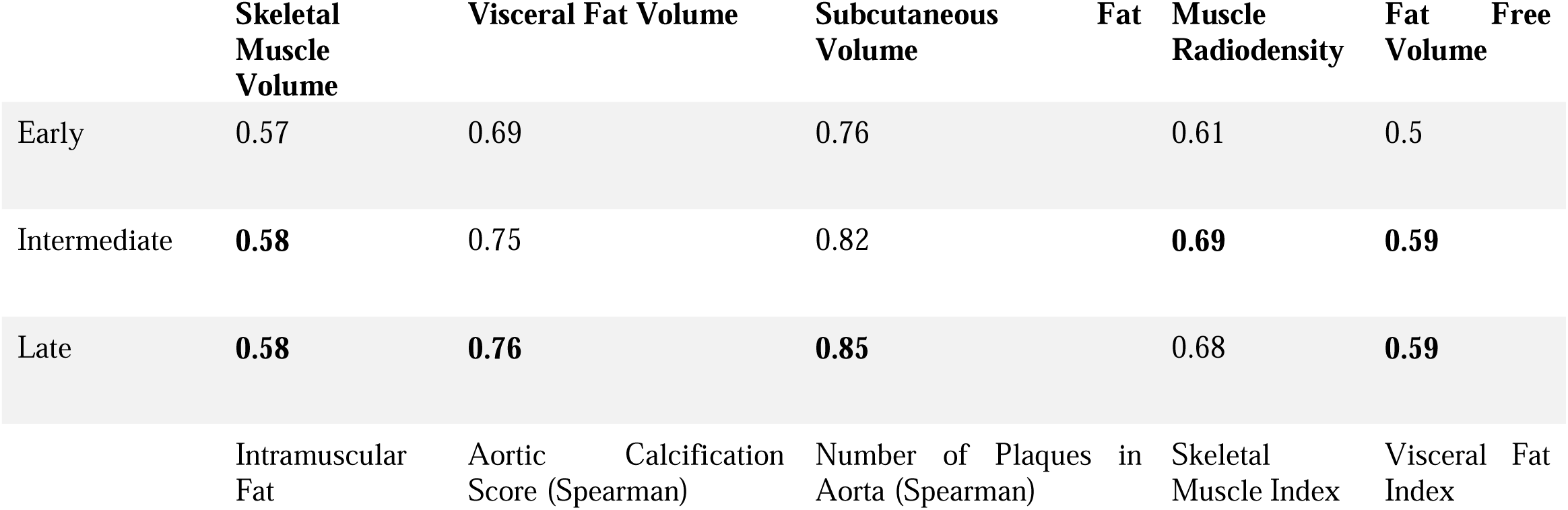
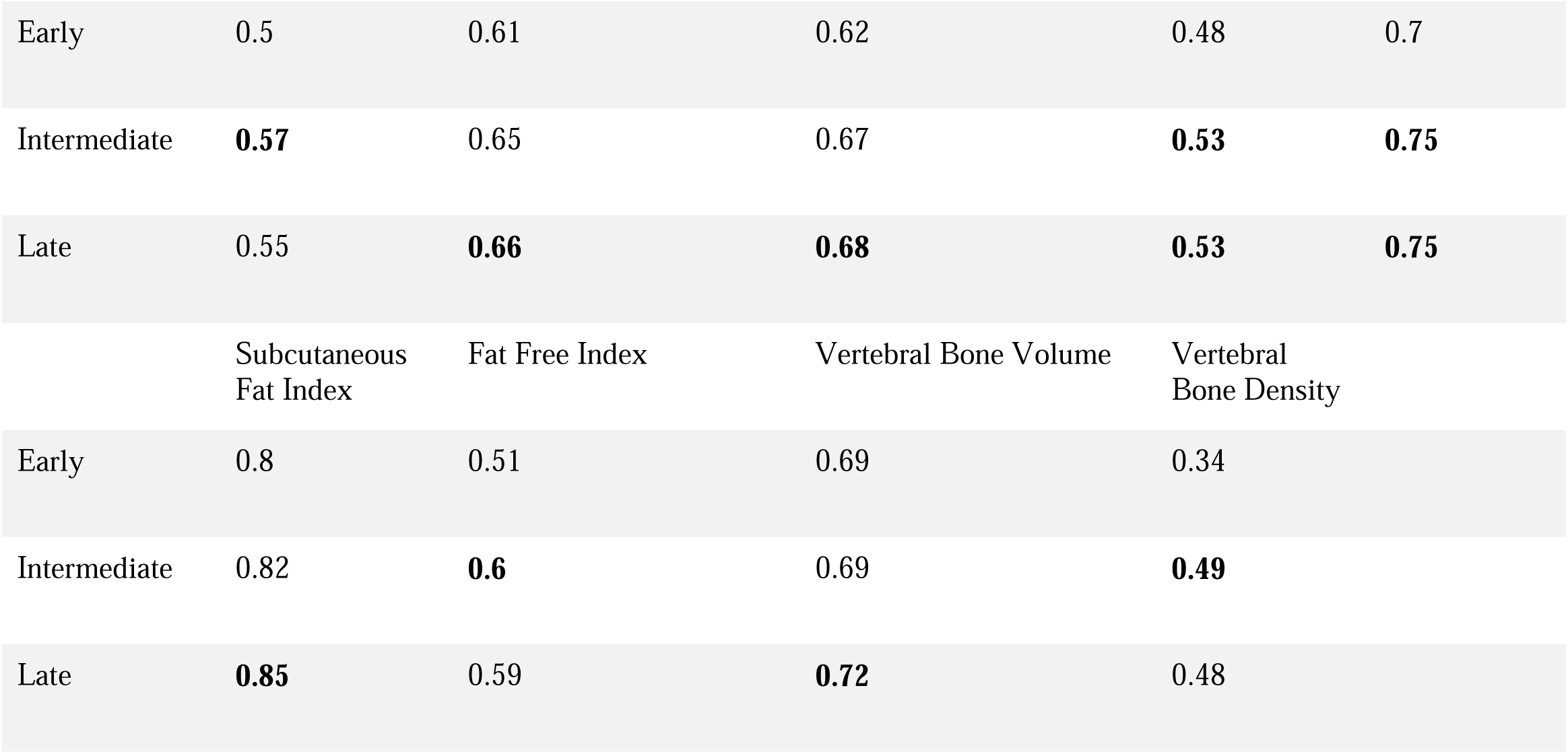
Test set performance across various fusion strategies. (Pearson’s Correlation)

### L3 Slice Level Models

Our L3 level model achieved similar performance in the top performing categories to the volumetric body composition (T12-L5) models. Its estimates on the hold out test set achieved a correlation of 0.81 (0.76 – 0.85) for subcutaneous fat area and 0.74 (0.67-0.80) for visceral fat area.

### Explainability

The aggregate explainability saliency maps for the prediction of subcutaneous fat volume, visceral fat volume and vertebral bone volume and skeletal muscle volume can be viewed in **figure 3**.

### Fairness Analysis

We investigated the performance of our model within different age, sex, and BMI subgroups of our test cohort to identify potential biases in our model. In general, although there was variability in model performance across various subgroups, no definitive trend is observable. The results for the top performing body composition metrics can be seen in **supplementary material Table S1**.

## Discussion

In this study we developed a multimodal deep learning model using chest radiographs and four clinical variables to estimate various body composition measures. Our model achieved good predictive performance in estimating subcutaneous adipose tissue, visceral adipose tissue, vertebral bone volume and to a lesser extent skeletal muscle volume. The resulting model can be used for a larger population of people to get estimated body composition metrics that are more accurate surrogates compared to weight and BMI alone while don’t involve costly imaging or large amounts of radiation.

Our multimodal model outperformed unimodal clinical-only and imaging-only models with performance increase being especially noticeable on estimating subcutaneous fat volume visceral fat volume and vertebral bone volume. The combined model did not perform noticeably well on tissue level body composition metrics or aortic calcification score levels. We also trained models to predict mid-L3 level body composition metrics. We showed that the models for mid-L3 body composition metrics perform similar to the volumetric ones.

Our clinical only model that included sex at birth, weight, height, and age could be used to estimate some body composition measures as well, especially subcutaneous fat area, skeletal muscle area and aortic calcification score. However, a clinical only model performs poorly when estimating other body composition metrics, especially visceral fat area. Visceral fat area is particularly important as it is correlated with adverse outcomes in various diseases (3,5).

The ability to differentiate subcutaneous and visceral fat is specifically important due to the importance of visceral fat volume in prediction of various risk factors from cardiometabolic disease to cancer (3,6). Another important body composition metric which is proven to be a risk factor for adverse outcomes is sarcopenia (21). We looked at both skeletal muscle volume, radiodensity and intramuscular fat in our study. While our final model only achieved moderate correlation in predicting skeletal muscle volume, during training and validation we observed more promising performance. This decline in performance could be attributed to overfitting of the model or could be due to other differences between the training and testing cohorts. In addition, our model had moderate to good performance in estimating the muscle radiodensity (r = 0.69, 0.61 – 0.75). Additional work is required to explore the possibility of estimating these two metrics from radiographs.

### Importance of fusion timing in final model performance

Previous studies that investigated the timing of fusion for medical data, have found conflicting answers. For example, study that looked into developing a multimodal model for the detection of pulmonary embolism concluded that the model that used late fusion performed best (17). Other studies, looking at fusion of various types of EHR data or imaging have found that intermediate fusion performed best for their study (22,23).

In our study, we found that while the performance of late and intermediate based fusion models was very close, the late fusion model had a higher performance. Interestingly, for the L3 level body composition metrics, the performance of the intermediate fusion model was higher. These differences show the importance of experimenting with different fusion strategies in multimodal deep learning studies. Various factors might be influencing the model performance across different fusion strategies. For example, in late fusion the imaging only and the clinical only models are trained and then their weights are frozen. This will lead to a simpler model at the end that is less likely to overfit to the training data due to the lower number of trainable parameters. This approach is specifically useful in cases in which the training sample size is small. On the other hand, early and intermediate fusion strategies allow the network to learn patterns across the various modalities.

### Model explainability, fairness and feature importance

The saliency maps in **Figure 4** show that for the estimation of subcutaneous and visceral fat tissue volume, the model looks at the mediastinum (which contains visceral adipose tissue), the suprasternal region and the sides of the trunk. Although the saliency maps overlap, they put different importance on different parts. For example. The visceral fat volume model is more focused on the mediastinum and neck region compared to the subcutaneous fat volume model. The model for vertebral bone volume focuses on the part of the image that contains the lower thoracic vertebrae behind the mediastinum. In general, our saliency maps show that the model is looking into the appropriate regions of the radiograph when estimating body composition metrics.

**Figure 4.**
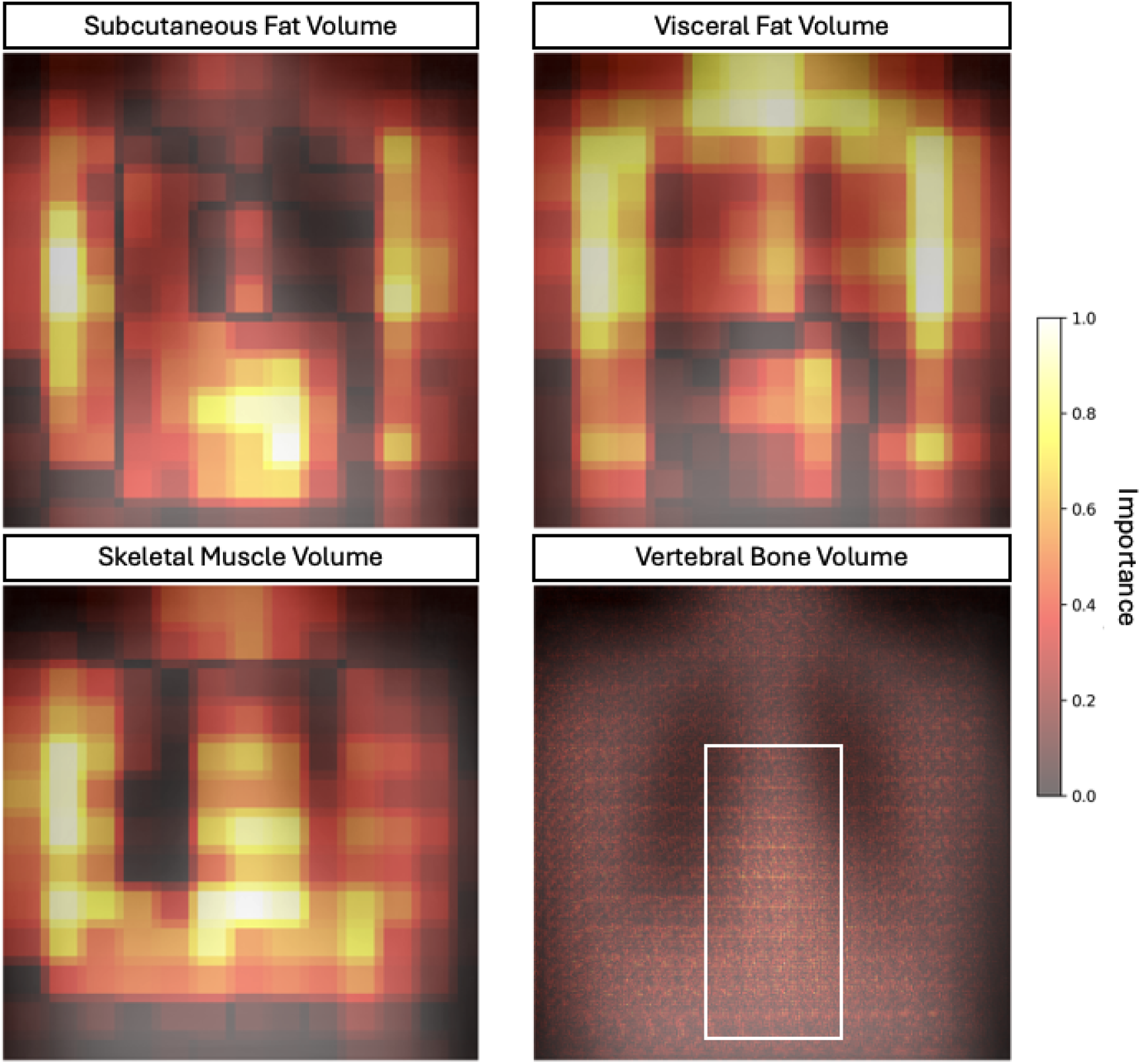
Saliency Maps for feature importance in prediction of the top performing body composition metrics. For the first three occlusion-sensitivity saliency map is depicted while for vertebral body volume. The images are aggregates of individual chest radiographs and their respective saliency maps in the hold-out test cohort.

Our fairness analysis showed that our model had variable performance across different age and sex groups. While these differences did not point to a definitive trend, it is important to note them when applying this model to other populations.

### Failure Analysis

Three cases that were among the worst performing in the test cohort can be seen in **Figure 5**. Compared to a typical chest radiograph, these demonstrate problems like incomplete coverage of the trunk on both sides, patient rotation, inadequate lung inflation and low exposure. Based on these findings, we hypothesize that our model’s performance could be higher if provided with high quality chest radiographs.

**Figure 5.**
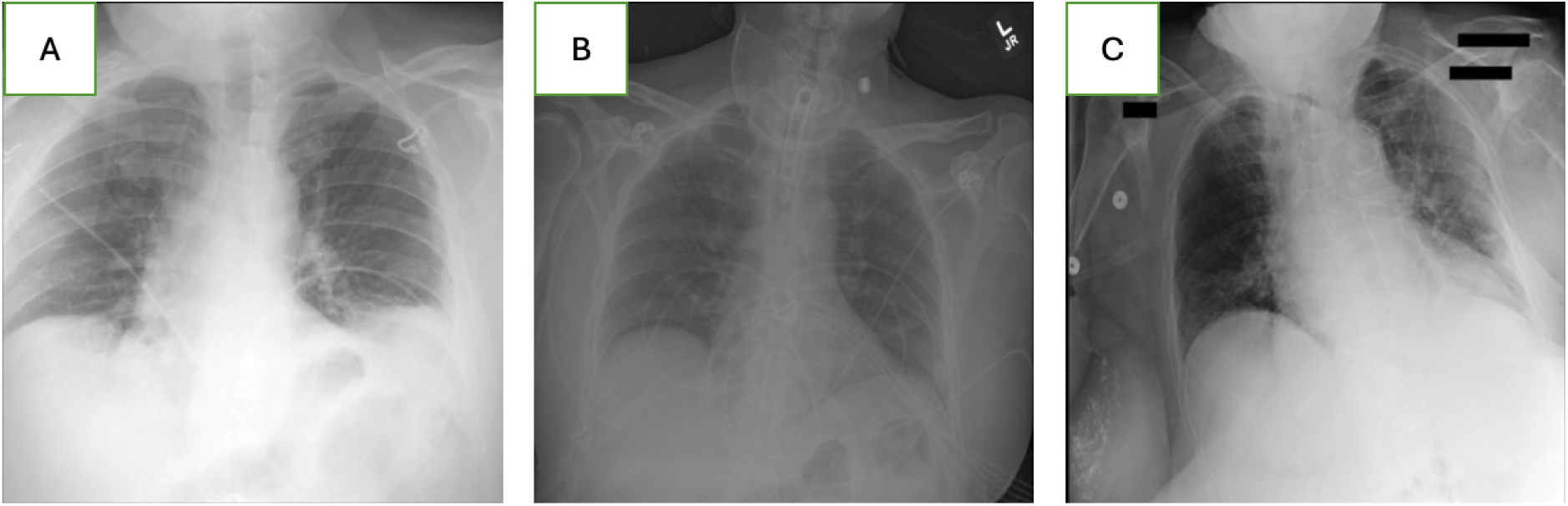
Three samples of the of the radiographs in which the model had poor performance. A. Parts of the chest radiograph that are important to the model are missing from the image. B. This chest radiograph is dark. C. The patient is rotated, and the lungs are not properly inflated.

### Limitations

We believe that although we achieved good performance on multiple tasks in this study, due to computational constraints and data loss, the sample size of our study was relatively small for this task. Additionally, the maximum 90 days difference between the chest radiograph and CT scan could potentially introduce noise into our data as body composition could change rapidly due to various factors like diet, exercise, or disease. Another limitation of our study is using abdominal CT scan-based body composition metrics. Finally, poor performance of the model on estimating Fat Free Index could therefore be also attributed to the random difference between train and test sets as was determined in our ANOVA analysis.

In conclusion, we developed a multitask multimodal deep learning model to estimate various body composition metrics, including subcutaneous fat volume, visceral fat volume and vertebral bone volume using a chest radiograph and four relevant clinical variables. We also explored how various strategies to fusion strategies can contribute to varying model performance.

## Data Availability

The data used in this study are available to all Truveta subscribers and may be accessed at studio.truveta.com.

