## Supplementary Material for "Multimodal Deep Learning Model to Estimate CT-based Body Composition Measures Using Chest radiographs and Clinical Data"

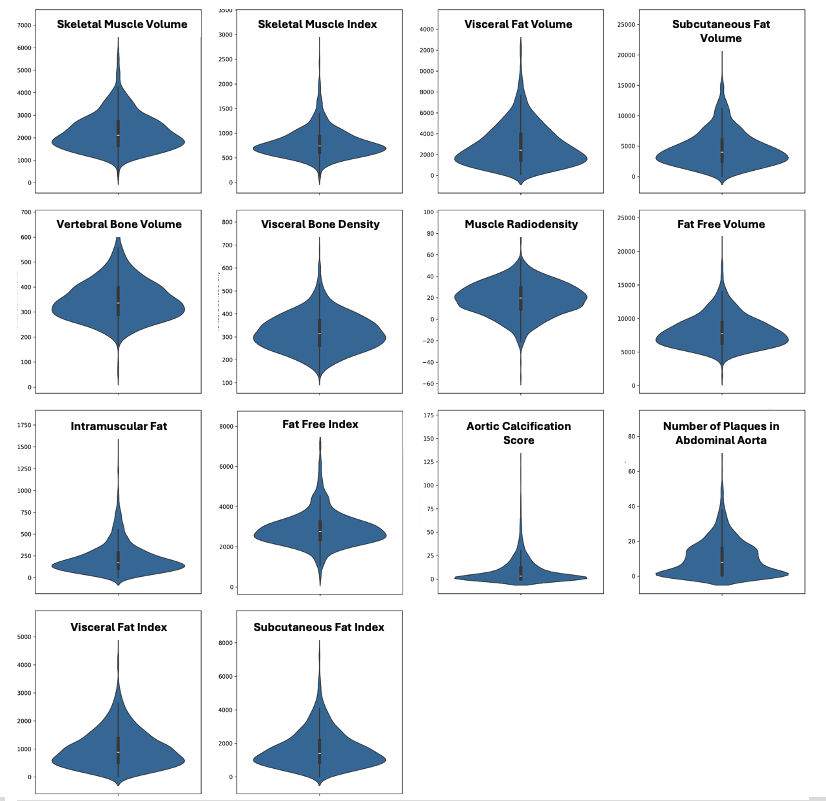

Figure S1 Distribution of body composition metrics calculated for our cohort.

Table S1: Distribution of mid-l3 level body composition metrics

|  | Train | | **Validation** | | **Test** | | **All** | | **Significance Test** |
| --- | --- | --- | --- | --- | --- | --- | --- | --- | --- |
|  | mean | std | mean | std | mean | std | mean | std | p-value |
| Skeletal Muscle Area (cm^2^) | 125.19 | 39.14 | 125.55 | 40.69 | 125.09 | 40.08 | 125.23 | 39.54 | 0.99 |
| Skeletal Muscle Index | 44.55 | 14.6 | 46.31 | 17.58 | 44.45 | 16.51 | 44.81 | 15.49 | 0.37 |
| Visceral Fat Area (cm^2^) | 166.41 | 112.71 | 171.21 | 102.83 | 166.66 | 111.93 | 167.23 | 110.96 | 0.87 |
| Visceral Fat Index | 58.81 | 39.98 | 62.06 | 35.12 | 59.11 | 40.12 | 59.39 | 39.26 | 0.61 |
| Subcutaneous Fat Area (cm^2^) | 238.02 | 149.96 | 255.73 | 142.55 | 237.12 | 138.73 | 240.67 | 146.71 | 0.33 |
| Subcutaneous Fat Index | 86.92 | 59.24 | 96.6 | 62.28 | 85.23 | 52.19 | 88.13 | 58.52 | 0.1 |
| Fat Free Area (cm^2^) | 369.48 | 96.71 | 379.41 | 124.86 | 377.9 | 111.37 | 372.69 | 104.55 | 0.38 |
| Fat Free Index | 132.08 | 37.74 | 140.52 | 53.04 | 134.85 | 45.96 | 133.96 | 42.24 | 0.05 |
| Intramuscular Fat Area (cm^2^) | 12.56 | 10.08 | 11.79 | 9.56 | 12.86 | 10.63 | 12.5 | 10.1 | 0.55 |
| Vertebral Bone Area (cm^2^) | 18.93 | 4.84 | 18.98 | 4.91 | 18.51 | 5.18 | 18.86 | 4.91 | 0.51 |
| Vertebral Bone Density (HU) | 326.38 | 83.42 | 329.01 | 74.68 | 333.66 | 88.05 | 328.21 | 82.99 | 0.52 |

Table S2: Model performance (AUROC) across different subgroups in the test set. While there is no general pattern, there is variability in model performance with respect to age and BMI.

|  | Subcutaneous Fat Volume | Visceral Fat Volume | Skeletal Muscle Volume | Vertebral Bone Volume |
| --- | --- | --- | --- | --- |
| Age 18 - 39 | 0.92 | 0.7 | 0.73 | 0.85 |
| Age 40 - 59 | 0.79 | 0.71 | 0.54 | 0.6 |
| Age 60 - 74 | 0.81 | 0.77 | 0.7 | 0.75 |
| Age > 74 | 0.81 | 0.78 | 0.48 | 0.67 |
| Female |  |  |  |  |
| Male | 0.8 | 0.74 | 0.55 | 0.45 |
| BMI <25 | 0.61 | 0.68 | 0.52 | 0.59 |
| BMI 25-30 | 0.63 | 0.62 | 0.57 | 0.7 |
| BMI 30-35 | 0.67 | 0.72 | 0.68 | 0.81 |
| BMI >35 | 0.83 | 0.68 | 0.54 | 0.75 |

**Supplemental Section 1, Data Acquisition:**

Truveta provides access to continuously updated and linked EHR and claims data including demographics, conditions, encounters, immunizations, medications, laboratory results, procedures, clinical notes, and images. Through syntactic normalization, similar data fields from different health care organizations are mapped to a common schema referred to as the Truveta Data Model (TDM). Once organized into common fields, the values are normalized to standard ontologies such as ICD-10, SNOMED-CT, LOINC, RxNorm, and CVX, through semantic normalization. The normalization process employs an expert-led, artificial intelligence driven process to accomplish high-confidence modeling at scale. De-identification is attested to through expert determination in accordance with the HIPAA Privacy Rule.

In case of patients with multiple imaging that fit the criteria, the last abdominal CT and the closest posteroanterior (PA) or anteroposterior (AP) chest radiography to the abdominal CT were selected. Weight was determined from the observation closest to the date of the radiography. In addition to height and weight, we determined the age at time of chest radiography and the sex at birth for all patients. We normalized all weights to kilograms and all heights to meters.

**Supplemental Section 2, Body Composition Calculation:**

Area was calculated by multiplying the number of pixels in each region of interest by the area of each axial pixel. Calcified plaques were identified using any contiguous voxels with a Hounsfield units (HU) value significantly larger than the median HU value of the aorta as described in the paper (13). Agatston scoring algorithm is then used to score the abdominal aorta calcification (14). Skeletal muscle fat volume was calculated by multiplying pixel area by the number of pixels in the skeletal muscle with a HU value in the range of adipose tissue (15). For abdominal aorta calcification score and number of calcified plaques only volumetric segmentation of abdominal aorta was used.

**Supplemental Section 3, Preprocessing:**

All retrieved radiographs were manually reviewed by a physician scientist to ensure only PA or AP chest x-rays are included and images contain no significant artifacts. Chest radiograph pixel values were normalized using z-score normalization. All x-rays were resized to 512*512 pixels. Z-score normalization was applied on all numerical variables and all calculated body composition metrics to ensure numerical stability during training using mean and standard deviation of the training set.

**Supplemental Section 3, Fusion Strategies:**

For early fusion, we used a fully connected layer to generate encodings for the clinical variables that were in turn added to the input image and passed through the rest of the network. For intermediate fusion, the encodings generated by the CNN were concatenated with the encodings generated by the shallow neural network and a final fully connected layer was added on the top. For late fusion, two separate imaging only and clinical only models were developed. The predictions of these two networks were collected and passed through a fully connected neural network.
